# User engagement in a digital health intervention designed for young people who have experienced technology-assisted sexual abuse (i-Minds trial)

**DOI:** 10.1101/2025.06.27.25330407

**Authors:** Sandra Bucci, Xiaolong Zhang, Kaja Dabrowska, Amanda Larkin, Ethel Quayle, Matthias Schwannauer, Filippo Varese, Pauline Whelan

**Author notes:** Corresponding author, Professor Sandra Bucci, 2nd Floor, Zochonis Building, Brunswick Street, Manchester M13 9PL, Manchester, UK.

## Abstract

**Background:** Technology-assisted sexual abuse (TASA) mostly involves the production and non-consensual sharing of sexual images; however, evidence-based support for young people (YP) who have experienced TASA is scant. Digital Health Interventions (DHIs) have the potential to increase access to support and provide timely therapeutic input in a familiar format to YP. However, studies describing engagement with DHIs is nascent.

**Objective:** To describe engagement patterns for young people who used the i-Minds app.

**Methods:** The i-Minds app is a co-designed mentalisation-based DHI for YP who have experienced TASA. Usage data was collected during the 6-week intervention window using Matomo analytics software and analysed according to the AMUsED framework.

**Results:** Forty-one participants were onboarded to the app. Of these, 95% completed the introductory mandatory module, and nearly half completed the remaining three modules. Median duration of app engagement was 33 days. Most participants used the app on weekdays in the afternoon. Demographic variables, namely gender not matching with sex assigned at birth/prefer not to disclose and higher baseline clinical severity were associated with higher app engagement.

**Conclusions:** Participants showed high module completeness and engagement duration, suggesting the potential for real-world use. Potential participant-level predictors of engagement, such as gender identity and severity of TASA related traumatic stress and emotional distress, were identified. Achieving satisfactory engagement in DHIs is challenging yet necessary for delivering effective interventions. Future studies should explore participant-level predictors of engagement to inform real-world use of DHIs with a diverse sample.

## Background

Using the internet has become a routine part of daily life for young people (YP), but it can place them at risk for various forms of harm. Technology-assisted sexual abuse (TASA) is one form of online harm that can occur across multiple digitally-enabled platforms and applications. TASA can involve YP being coerced into sharing sexual images of themselves, taking part in sexual activities via webcam or smartphone, or ‘sexting’; these behaviours can bring unique social and psychological harms (1)(2). Although multiple factors are likely involved in vulnerability to being exposed to TASA, the ability to accurately estimate others’ intentions and motivations (termed mentalising) when engaging in online spaces can be compromised because the usual assumptions based on verbal and non-verbal cues in real life interactions are more opaque when communication takes place online. This in turn can compromise a YP’s ability to evaluate risk and accurately mentalise (3). Digital health interventions (DHIs) have been shown to be a safe and acceptable way to support YP; they are unconstrained by location and time and can facilitate access to intervention strategies when traditional means of support are not available. In addition to understanding how feasible, acceptable, safe, and effective DHIs are with vulnerable YP, it is also important to understand engagement patterns, including how DHIs are accessed, used and the way users interact with a DHI. This can help researchers understand what can be done to improve the user experience and how to make interventions more effective. Engagement with DHIs is associated with therapeutic gains (4) but has been noted as a significant challenge in DHIs.

Reporting detailed usage analyses of DHI trials is relatively scant in the digital health literature (5), possibly because engagement with an intervention is a complex, multidimensional concept (6). Hassan and colleagues (7) argue that usage analyses of DHIs should not stop at the point of analysis of participant drop-outs or withdrawals but should explore engagement because DHIs require some level of ongoing effort from users, therefore rendering it an important factor to consider for an effective DHI to achieve its intended outcomes. Several methods are available for investigating engagement, including administration of self-reported questionnaires which can provide insights into attention, interest, satisfaction and subjective adherence with the intervention (8). Objective measures of engagement and how a DHI is utilised can be measured from embedded analytics and server logs, which can provide objective insights into overall usage and content-specific usage, factors that are valuable for optimising future DHIs. Usage can be operationalised with various analytical indicators, such as number or frequency of recording measures, number of features accessed, number of logins/sessions, and time spent using the app, to name a few (9); measures can be further grouped by frequency (e.g. number of logins), duration (e.g. total number of minutes logged in), breadth (e.g. number of pages) and depths (e.g. number of modules completed) (6).

i-Minds is a co-designed, theory-driven DHI underpinned by mentalisation principles, designed for YP (12-18 years) who have experienced TASA. Details of the development of the app, our method of co-design, subjective measures of user satisfaction and engagement, and findings from the trial are published elsewhere (10). In summary, we conducted a mixed-methods, non-randomised clinical feasibility study with 46 YP aged 12-18 years exposed to TASA across two UK sites, with a nested qualitative study to understand issues to be addressed in advance of conducting a powered efficacy trial (10). We found that it was possible to recruit and retain participants in a DHI trial of TASA, and that the i-Minds app was safe, acceptable and associated with promising signals of efficacy on key outcomes. User feedback indicated that participants generally had a positive experience using the app, positively increasing their knowledge/understanding of their own mental health and their motivation to address their mental health difficulties. In the current study, we report patterns of engagement among participants who took part in the i-Minds trial over the 6-week intervention window guided by the Analysing and Measuring Usage and Engagement Data (AMUsED) framework (5). The framework is used to guide researchers in familiarisation of the intervention, identification of key measures, specification of research questions, and preparation of data for analysis.

## Objective

Guided by the AMUsED framework with app usage data collected, we aimed to explore user engagement with the i-Minds app during the i-Minds non-randomised clinical feasibility trial. We specifically aimed to explore: i) content-specific completeness of the i-Minds app; ii) the timing and duration of engagement; and iii) association between baseline characteristics and app engagement.

## Methods

### Research design

The methods and results of the i-Minds non-randomised clinical feasibility trial regarding clinical outcomes are reported in the trial protocol (10) and outcome paper (*under review*). This paper focuses on the methods and results relevant to examining engagement patterns during the i-Minds clinical feasibility trial. The pre-registered trial (ISRCTN43130832) was approved by the National Research Ethics Committee (REC) of West Scotland REC 4 (approval number 22/WS/0083) and overseen by an independent oversight committee. All participants gave informed consent.

### Participants

Participants were recruited from Child and Adolescent Mental Health Services (CAMHS) in Edinburgh and Greater Manchester, one sexual assault referral centre (SARC) in Greater Manchester, and an NHS-commissioned e-therapy provider. Further details of the recruitment procedure are reported elsewhere (10). Potential participants were eligible to take part in the clinical feasibility trial if they were: i) aged 12-18 years; ii) exposed to TASA and report associated distress; iii) receiving support from CAMHS, SARC or an e-therapy provider and continued to be actively supported by services for the trial duration; iv) willing to use an app about TASA; v) proficient in speaking and writing in English; vi) capacity to consent; and, for e-therapy provider users only, willing to provide their username. YP with insufficient command of English, moderate learning difficulties, or at risk of current or recent suicidality (assessed by referrers) were excluded. Following recruitment, baseline assessments were completed to collect data about participant demographics and clinical characteristics. Participants could use their own smartphone and download the i-Minds app with support from a researcher (if requested) or they could opt to borrow a smartphone with the app preloaded. We asked participants to return the loaned handset at study end. Data network charges were paid for the intervention period, as was completion of research assessments.

### Intervention

The i-Minds app is a modular DHI available over 6-weeks. The app is designed to support help-seeking YP-OSA (i.e. those already under the care of services or an e-therapy provider) better mentalise in the online environment and is intended to be used as a stand-alone app. App content is organized into four modules: 1) mentalisation (mandatory module to complete before access to other modules is permitted); 2) psychoeducation (technology, relationships, sexual experiences); 3) emotional and mental health; and 4) trauma (focusing on the psychological and emotional consequences of trauma, including recognising and responding to ‘triggers’, addressing and improving feelings of guilt and shame, and reducing maladaptive avoidance). In addition to module content, participants can access a repository of multi-media material designed to support learning and promote engagement, including videos and audio exercises, a diary function, links to podcasts, blog posts and other useful links, real-life stories of recovery, emergency and safeguarding contacts, relaxing/breathing/mindfulness exercises, soothing visual images, and inspiring quotes. Topics areas are visually represented and navigation through the app uses a tree motif; the tree grows leaves as segments and modules are completed. Users are sent a daily prompt in the form of a standard app notification (at 5 pm), designed to prompt engagement, with a further notification three hours later (at 8 pm) if the initial prompt did not result in the use of the app; users can also self-initiate use at any time. Screenshots of the i-Minds app modules are shown in Supplementary Figure 1.

### Analysis

The AMUsED framework was used to systematically guide analysis of usage data (5). The AMUsED checklists were completed prior to analysis (see Supplementary Table 1). Stage 1 AMUsED checklist was used to specify specific workflow and content of the intervention, review data accrued, and understand the underlying mechanism of the intervention. Stage 2 AMUsED checklist was used to review usage metrics and available baseline characteristics, how they could be associated and to develop the research questions. Stage 3 AMUsED checklist was used to understand the data format, specify and plan tasks in order to prepare the data for analysis, and review available tools for analysis.

App usage data was collected using Matomo analytics, an open analytics platform (11). Matomo analytics collects a large variety of usage indicators, including user-specific measures such as time spent using the app (daily, overall), and number of visits and actions (daily, overall), as well as aggregate measures, such as total time spent using the app by all users during each day of the week or each time of day.

Descriptive statistics were used to characterise engagement frequency, duration and type of participants. The associations between baseline demographic and clinical characteristics and app engagement were assessed using Spearman’s correlation (continuous variables), independent t-tests (binary variables), or ANOVA (categorical variables). Due to small sample sizes in certain demographic categories, variables were regrouped to enable meaningful comparisons (i.e. gender matches sex assigned at birth, level of education, employment status, relationship status), while those that could not be regrouped or still had a small sample size after regrouping were not analysed (i.e. ethnicity, living status). Analyses were performed in R (version 4.3.2) (12). The P value threshold was set to 0.05.

## Results

### Sample characteristics

Of the 147 young people screened for eligibility, 72 referrals were made to the trial and 65 participants were deemed eligible; 46 young people consented to take part in the i-Minds clinical feasibility trial. Three participants withdrew consent prior to completing baseline assessment, leaving 43 participants completing baseline assessment and 41 participants onboarded to the app. Demographic and clinical characteristics of the 41 young people onboarded to the i-Minds app are reported in Table 1 (demographics of the wider i-Minds sample are reported elsewhere; *Bucci et al., under review*). The average age of participants was 15.2 (SD = 1.45) years. Most identified as female (70.73%) and White British (97.56%). A notable percentage of participants (14.64%) identified as non-binary/third gender or preferred not to disclose their gender identity, and 21.95% reported their gender did not match their sex assigned at birth or preferred not to disclose their gender. Most participants were living with parents (95.12%), currently a student (85.37%), single (78.05%) and did not use apps to help with their mental health (87.8%). Two-fifths (39.02%) of participants were taking medication, and 60.98% were receiving psychological support, with just over half having received a psychiatric diagnosis. As per eligibility criteria, all participants reported a history of distress associated with TASA. Most participants accessed the i-Minds app via their own phone; 4.88% (n=2) of the sample requested a study handset.

**Table 1.** Baseline Characteristics and service use (N=41).

| Characteristic |  | N (%) |
| --- | --- | --- |
| **Age (years)** | Mean (min-max) | 15.2 (12 – 18) |
| **Age group** | 12–14 | 12 (29.27%) |
|  | 15–18 | 29 (70.73%) |
| **Gender** | Man / Male (n/%) | 6 (14.63%) |
|  | Woman / Female (n/%) | 29 (70.73%) |
|  | Non-binary / Third gender | 3 (7.32%) |
|  | I would rather not say / unsure | 3 (7.32%) |
| **Gender matches sex assigned at birth** | Yes | 32 (78.05%) |
|  | No | 8 (19.51%) |
|  | Prefer not to say | 1 (2.44%) |
| **Ethnicity** | White British | 40 (97.56%) |
|  | Asian/Asian British | 1 (2.44%) |
| **Level of education** | Primary school | 28 (68.29%) |
|  | Secondary school | 12 (29.27%) |
|  | Further education | 1 (2.44%) |
| **Living status** | Living with parents | 39 (95.12%) |
|  | Living alone | 1 (2.44%) |
|  | Other | 1 (2.44%) |
| **Carer status** | Yes | 6 (14.63%) |
|  | No | 35 (85.37%) |
| **Employment status** | Student | 35 (85.37%) |
|  | Part time / casual job | 3 (7.32%) |
|  | Voluntary job | 3 (7.32%) |
|  | Other | 1 (2.44%) |
| **Relationship status** | Single | 32 (78.05%) |
|  | Partnered | 8 (19.51%) |
|  | Rather not say | 1 (2.44%) |
| **Study phone** | Own phone | 39 (95.12%) |
|  | Borrowed phone | 2 (4.88%) |
| **Deprivation Index** | Range | Scotland: 248 – 6,933<br>England: 4,282 – 32,503 |
| **Baseline service use and use of internet / being online** |  | N (%) |
| **Currently supported by CAMHS** | No | 2 (4.88%) |
|  | CAMHS / Child & Adolescent Mental Health Team | 32 (78.05%) |
|  | Sexual Assault Referral Centre (SARC) | 2 (4.88%) |
|  | e-therapy provider | 2 (4.88%) |
| **Use apps to support mental health** | Yes | 5 (12.2%) |
|  | No | 36 (87.8%) |
| **Have been in psychiatric hospital / ward** | Yes | 3 (7.32%) |
|  | No | 38 (92.68%) |
| **Take medications** | Yes | 16 (39.02%) |
|  | No | 25 (60.98%) |
| **Receive psychological support** | Yes | 25 (60.98%) |
|  | No | 16 (39.02%) |
| **Have received a diagnosis** | Yes | 22 (53.66%) |
|  | No | 19 (46.34%) |

### Engagement with i-Minds features

Of the 41 YP onboarded to the app, 95% users (39/41) completed the mandatory mentalisation module. For the three remaining modules, the trauma module had the most completions (46% (19/41) users completed the module), followed by the psychoeducation and emotional and mental health modules (39% (16/41) users completed each module). Additionally, 32%, 44%, and 34% of users accessed (but did not complete) the trauma, psychoeducation, and emotional and mental health modules, respectively. A breakdown of completion status of each module is shown in Figure 1. The total time spent on each module, multi-media resources, and diary is displayed in Supplementary Figure 2. Of the four intervention modules, the median total time spent on the mandatory mentalisation module was the lowest (2.48 minutes, IQR 1.03–4.87, range 0.05–22.15), whereas the median total time spent on the trauma module was the highest (7.87 minutes, IQR 3.98–12.66, range 0.08–56.22). Besides the intervention modules, the median total time spent on the multi-media resources and diary was 4.67 minutes (IQR 2.27–13.95, range 0.02–27.32) and 1.68 minutes (IQR 0.81–4.22, range 0–15.52), respectively.

**Figure 1.**
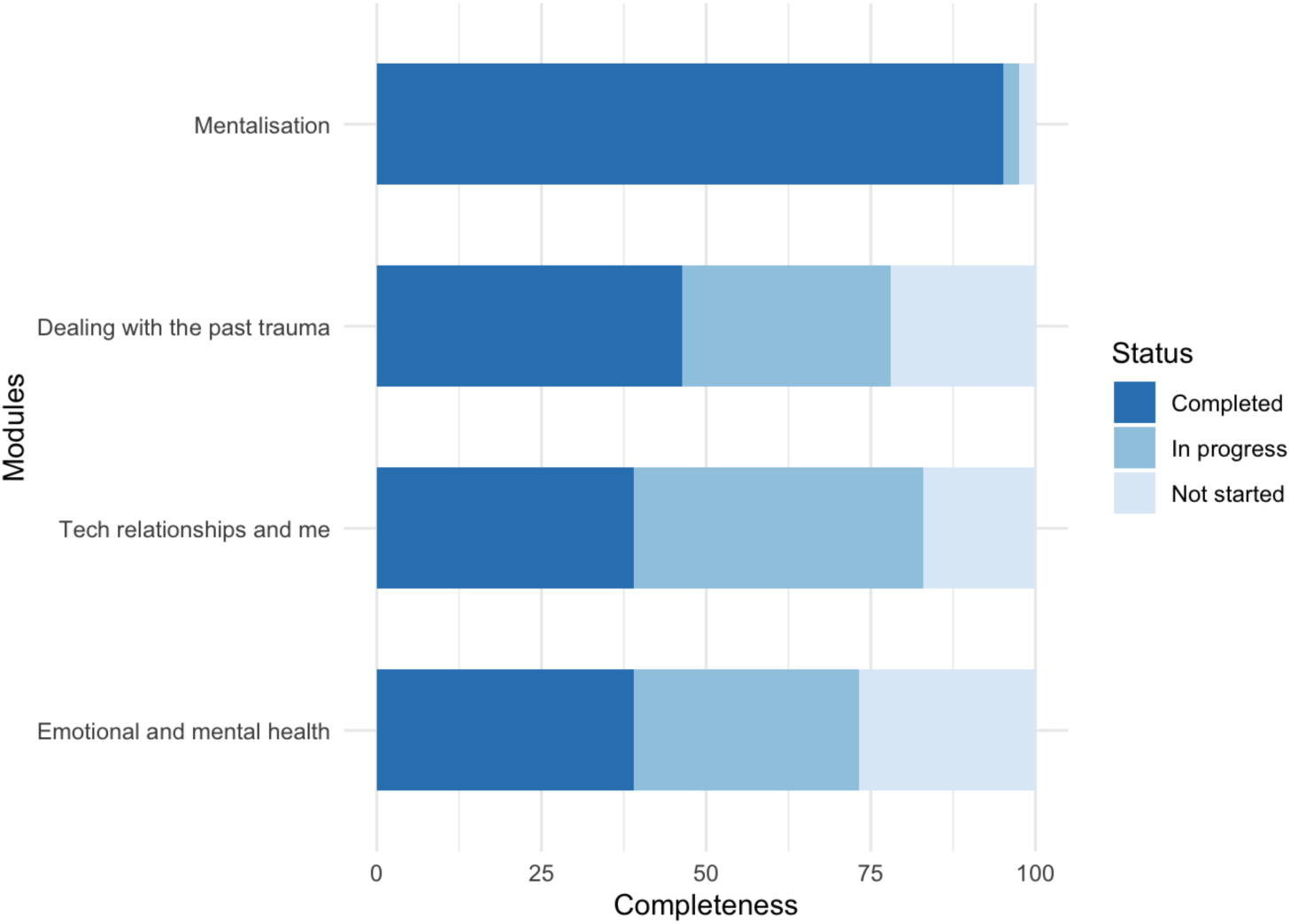
The completeness of the i-Minds modules

### Duration and timing of engagement

Figure 2 shows the probability of engagement with the app across six weeks. The median survival time was 33 days (95% CI 25–38). The distribution of daily total time spent on the app in each week is shown in Supplementary Figure 3. The median total time spent on the app ranged from 1.1 minutes (IQR 0.27– 5.43, range 0–34.15) in week 3 to 5.78 minutes (IQR 1.22–14.53, range 0–88.52) in week 1. Most participants spent relatively less time on the app compared to the median time spent, whilst a small number of participants spent notably longer using the app compared to their peers.

**Figure 2.**
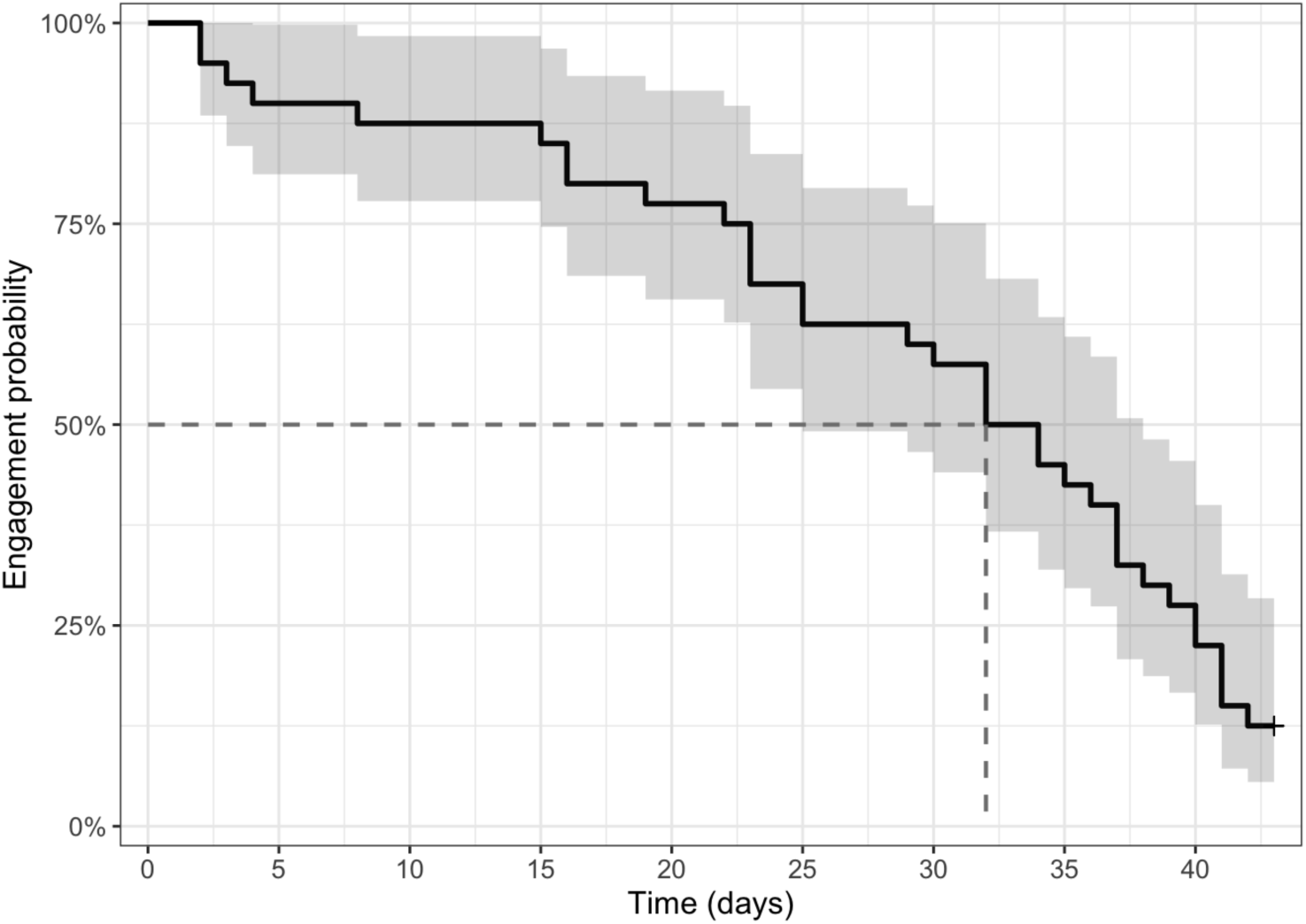
Kaplan-Meier plot of engagement over six weeks

Figure 3 shows the average time spent per user in a day. Participants mostly interacted with the app in the afternoon; 5pm was the highest use period in line with the app prompt schedule. As shown in Figure 4, participants spent more time on the app on weekdays than weekends; more specifically, participants spent most of time on the app on Thursday, followed by Wednesday and Monday.

**Figure 3.**
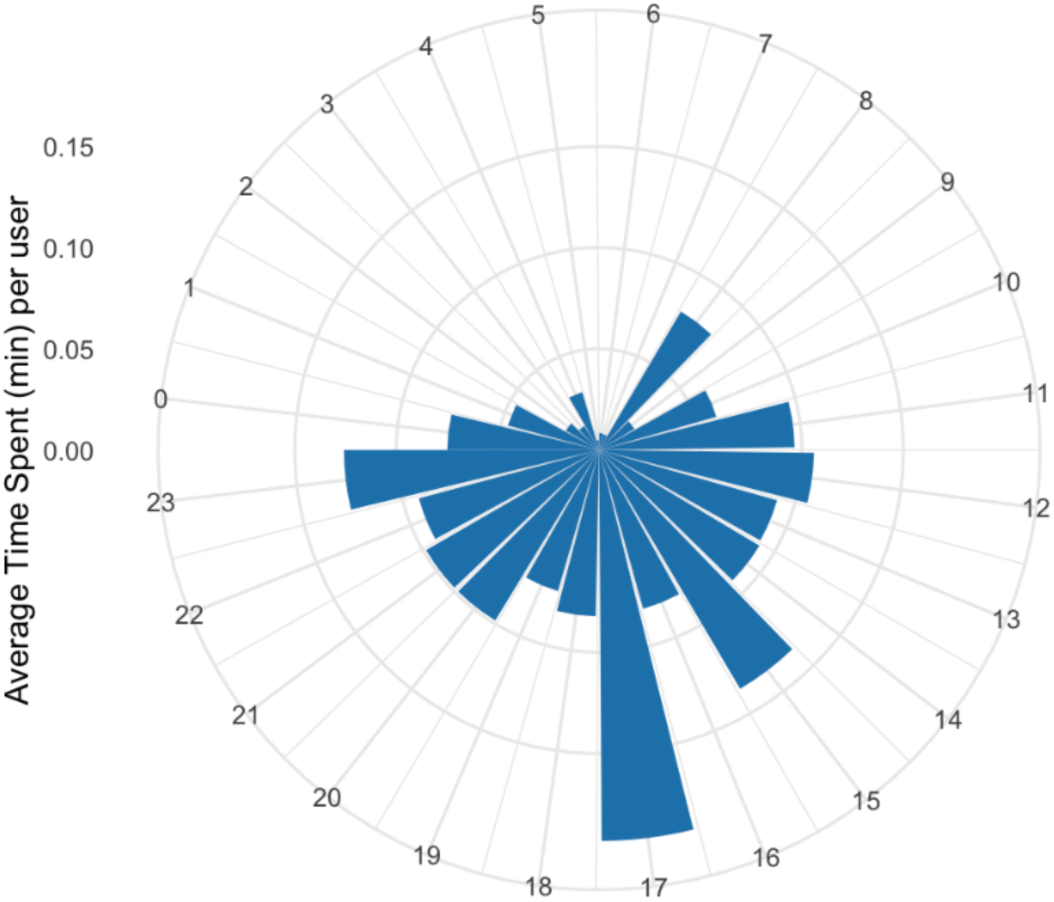
Average time spent using the app in a day

**Figure 4.**
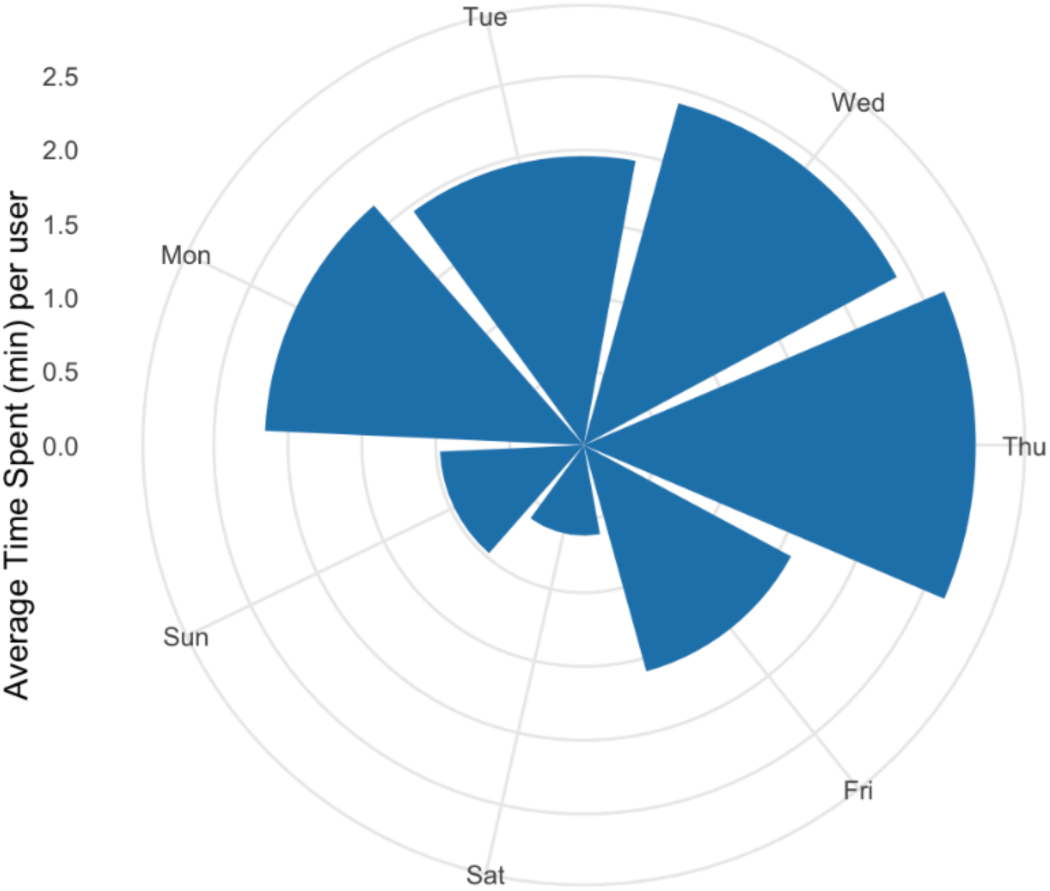
Average time spent using the app in a week

### Factors associated with engagement

We conducted correlation analysis of baseline demographic and clinical characteristics and engagement data. Among the demographic factors, participants who identified their gender as not matching with the sex assigned at birth or preferred not to disclose spent more time on the app (t=-4.83, p<0.001); no significant associations were found between other demographic features and app engagement: age (rho=0.07, p=0.664), gender (F=2.36, p=0.087), level of education (t=-0.11, p=0.91), carer status (t=0.85, p=0.401), employment status (t=0.01, p=0.993), and relationship status (t=-0.21, p=0.834). Of the baseline clinical assessments, longer total time spent on the app was associated with a higher baseline intrusion score (rho=0.33, p<0.05) on the Child Revised Impact of Events Scale (CRIES) (13), and anxiety (rho=0.36, p=0.025), depression (rho=0.39, p=0.015) and total emotional distress scores (rho=0.36, p=0.026) of the Revised Child Anxiety and Depression Scale–25 item version (RCADS–25) (14).

## Discussion

### Main findings

This study aimed to understand usage patterns of the i-Minds app designed for YP who have experienced distress associated with TASA in a clinical feasibility trial. We found that, of the four intervention modules, 95% participants onboarded to the app completed the mandatory mentalisation module, and the remaining three modules were completed by nearly half of the participants. Survival analysis of app disengagement indicated that the median survival time was 33 days. Most participants used the app in the afternoon and spent more time on the app on weekdays than weekends. Correlation analysis showed that participants who reported their gender identity differing from their sex assigned at birth/preferred not to disclose, higher levels of intrusion and emotional distress of TASA at baseline spent more time using the app.

Engagement of DHIs in real-world settings remains limited, and the reduction of usage over time was commonly reported (15)(16). As the nature of intervention and definition of engagement varies across studies, a direct comparison of engagement is difficult. A recent systematic review on engagement of smartphone-delivered interventions found that the mean percentage of participants completing the designed intervention content was between 34% and 64% for a range of mental health conditions, including depression, anxiety, stress, and general mental health (15). Another systematic review on YP’s engagement with DHIs in general showed that the completion rates of intervention content varied hugely across studies, ranging from 3.6% to 100% (17). Considering we did not incentivise participants to engage with the app, i-Minds demonstrated relatively high engagement compared to other DHIs by successfully achieving 95% of participants completing the mandatory module and nearly half of the participants completing the remaining three modules. Moreover, the trajectory of engagement with i-Minds declined steadily, whereas some DHIs trials showed a drastic engagement reduction over time (18)(19).

Identifying an approach to record and report on usage/engagement data for DHIs can be complex, as engagement is a multidimensional concept that includes behavioural, cognitive, and emotional components (20). However, many studies of usage/engagement of DHIs do not report these in a systematic manner, despite frameworks proposed by researchers to investigate engagement and its impact (21)(22)(23). This may be due to the lack of implementing appropriate tools to facilitate the systematic identification, extraction, and analysis of usage data (5). To address this issue, we employed the AMUsED framework to support a transparent approach to analysis and reporting. The AMUsED framework contains comprehensive checklists establishing a systematic guide for analysing usage data by drawing together potential measures of usage and identifying which are meaningful to the intervention, generating specific research questions to act as testable hypotheses, and supporting data preparation and selection of methods for analysis (5). Additionally, we used Matomo to collect objective data to support the evaluation of app usage; using a contemporary analytics software tool enabled us to collect a huge volume of detailed usage and engagement user data, which is one of the significant advantages of such tools.

Understanding the relationship between engagement and mental health outcomes is a critical step for optimising DHIs to be used more effectively. A recent meta-analysis showed that more frequent engagement with DHIs was associated with better mental health outcomes (4). However, due to the non-randomised study design, we were not able to determine the ‘effective dose’ of the i-Minds app (i.e. how much ‘engagement’ with the app is sufficient to bring about clinical change). Instead, we found app engagement was significantly associated with participants’ gender identity and the level of distress caused by TASA at baseline. This might support the idea that, when app content is relevant to the user, engagement increases (24)(25). A future RCT is warranted to further explore the relationships between engagement with i-Minds and its potential clinical benefits.

We did not use the usage and engagement analytics in real-time to support engagement (e.g. to identify when a participant had stopped engaging with the app and to encourage them to re-engage). However, in future, the app could provide a nudge to the YP if engagement drops below pre-defined/agreed threshold or engagement analytics could be used to invoke human support (e.g. call from supporting therapist).Our analysis of usage/engagement data can help us understand how digital tools may reach groups that may have traditionally been disadvantaged from accessing mental healthcare and as such may therefore have potential to address and reconfigure existing health inequalities.

### Strengths and limitations

This study explored objective and detailed usage data from the i-Minds app to characterize engagement over time. We did not define *a priori* hypotheses or pre-specified thresholds for engagement; the study was exploratory in nature. As the sample was largely White British help-seeking females, it is not clear how transferable findings around engagement with a DHI of this nature are to other groups of YP who have experienced TASA. Clinicians made decisions about YP they would refer to the i-Minds trial.

Decisions were based on factors such as whether a young person was well and ready enough to take part, or whether the DHI ‘fit’ with the treatment model they were using. It is possible that the trial attracted more digitally literate YP with more positive attitudes towards DHIs than the broader population of YP who have experienced TASA. Furthermore, usage analysis is based on automatically collected analytics from the app. Engagement is a complex concept that ideally requires integrating adjacent constructs, including predictors, behavioural outcomes and other measures, such as attention, affect and interest. Nonetheless, usage data analysis is still valuable to explore how the intervention was utilised, which was the aim of this study. However, the results should be interpreted with caution as inactive usage may exist and lead to an overestimation of engagement. While certain aspects of the i-Minds trial were designed to reflect real-world conditions, such as lack of financial incentives for app usage, others, such as paid network charges for the intervention window and provision of phones and data plans, may limit generalisability. Finally, the measures administered in the i-Minds trial, though not included in analyses reported here, might have influenced engagement with the app.

## Conclusions

There are currently no evidence-based assessments or interventions for YP exposed to TASA. Achieving satisfactory engagement in DHIs is challenging yet necessary for delivering effective interventions. This study analysed usage data from the co-designed, mentalisation-based DHI with YP who have experienced TASA to provide a detailed analysis of patterns of engagement and attrition. We have demonstrated that a sizeable proportion of participants used the app for the duration of the study, suggesting the potential for real-world implementation of the i-Minds app. Gender identity differing from sex assigned at birth, higher baseline intrusion and emotional distress related to TASA were identified as factors associated with higher engagement levels. Future studies should further explore the relationships between baseline demographic and clinical characteristics and DHIs engagement and take advantage of opportunities to exploit detailed usage data to inform the real-world implementation of DHIs with diverse samples in practice.

## Acknowledgments

The authors would like to thank the participants who took part in the i-Minds clinical feasibility trial, the healthcare professionals and researchers who supported the study, and the advisory groups who provided valuable insights and input throughout the broader programme of work. The authors declare that the research was conducted in the absence of any commercial or financial relationships that could be construed as a potential conflict of interest. This study is funded by the National Institute for Health and Care Research Health and Social Care Delivery (NIHR HSDR) programme (NIHR131848). The relevant Institutional and Health Research Authority ethics approvals were granted (REC Number: 21/WS/0160).

## Data Availability

The data set generated in this study is not publicly available owing to ethical restrictions for personal health information; however, limited, deidentified data may be made available from the corresponding author on reasonable request.

## Authors’ Contributions

SB, XZ, and KD drafted the manuscript. XZ conducted the data analysis. SB, FV, MS, and EQ conceptualised the study, secured funding, and led the development of the i-Minds app. MM is responsible for engineering the i-Minds app. PW led the participatory design work. SB, EQ, MS, FV and AL supervised data collection. All authors critically revised the manuscript for intellectual content and approved the final version of this protocol.

---

**Supplementary Table 1.**
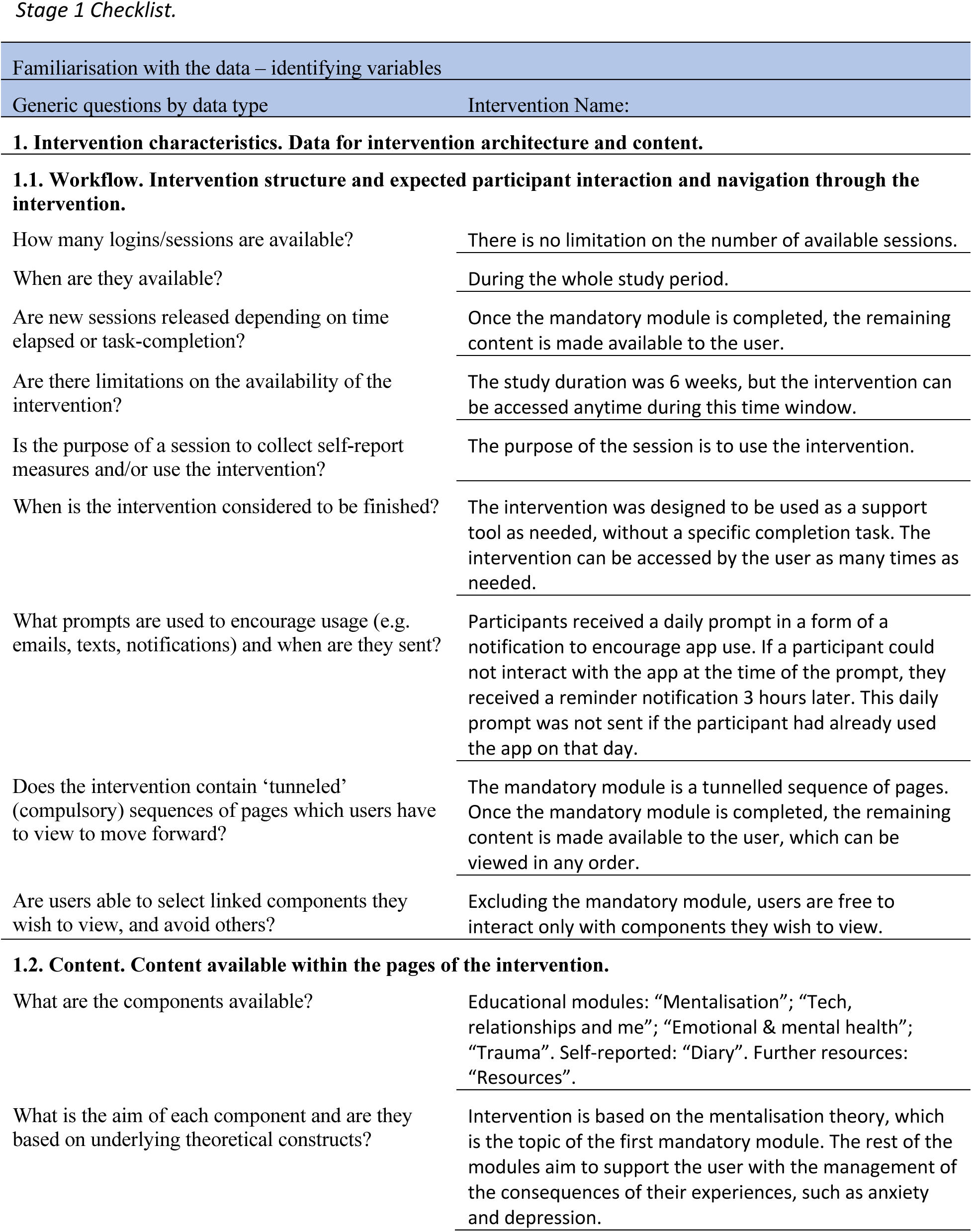
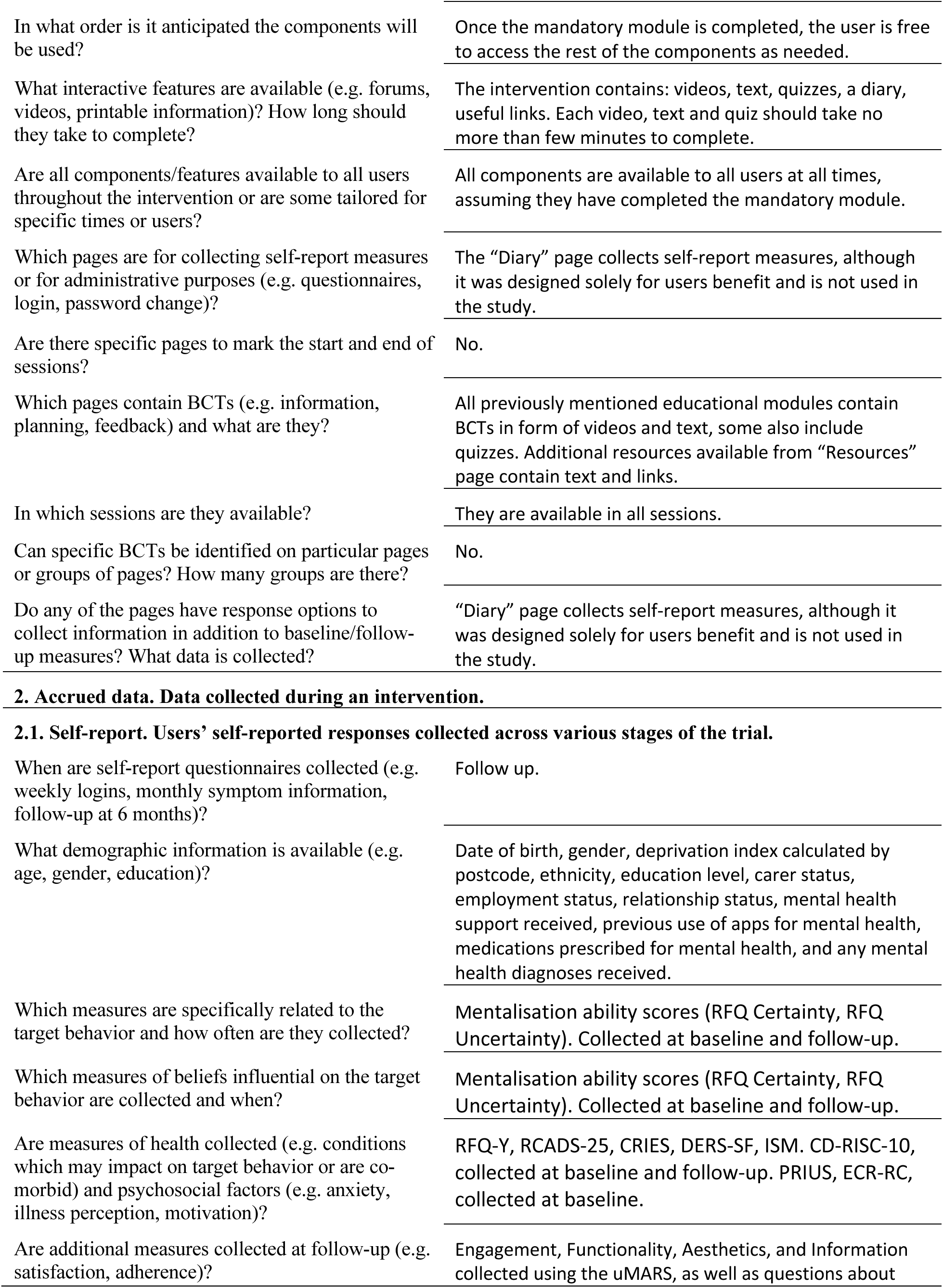
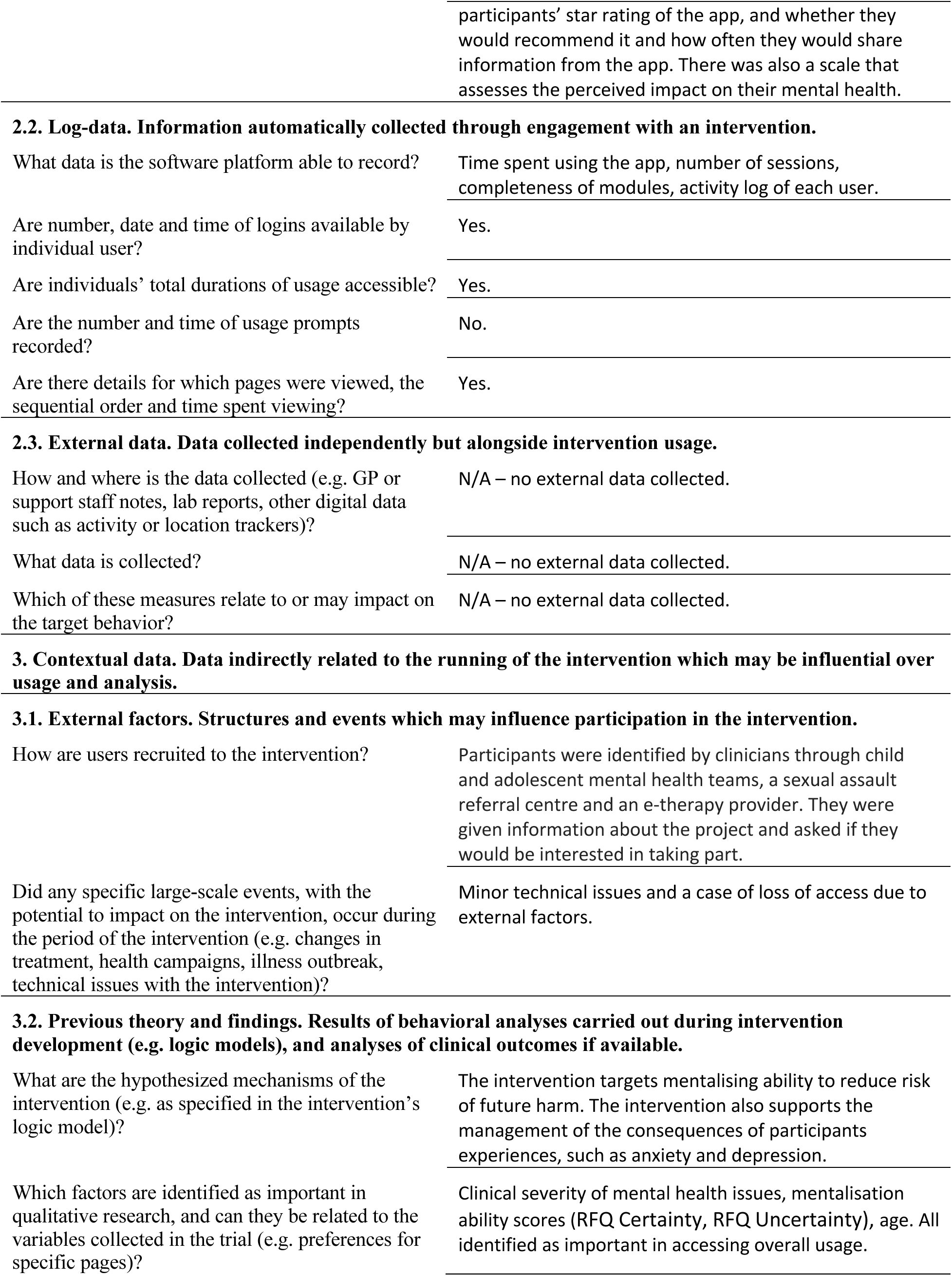
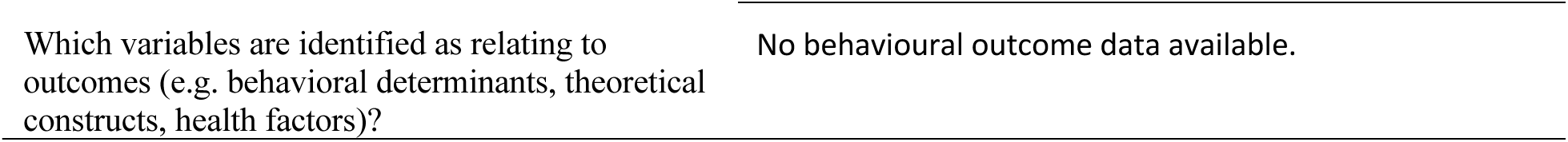
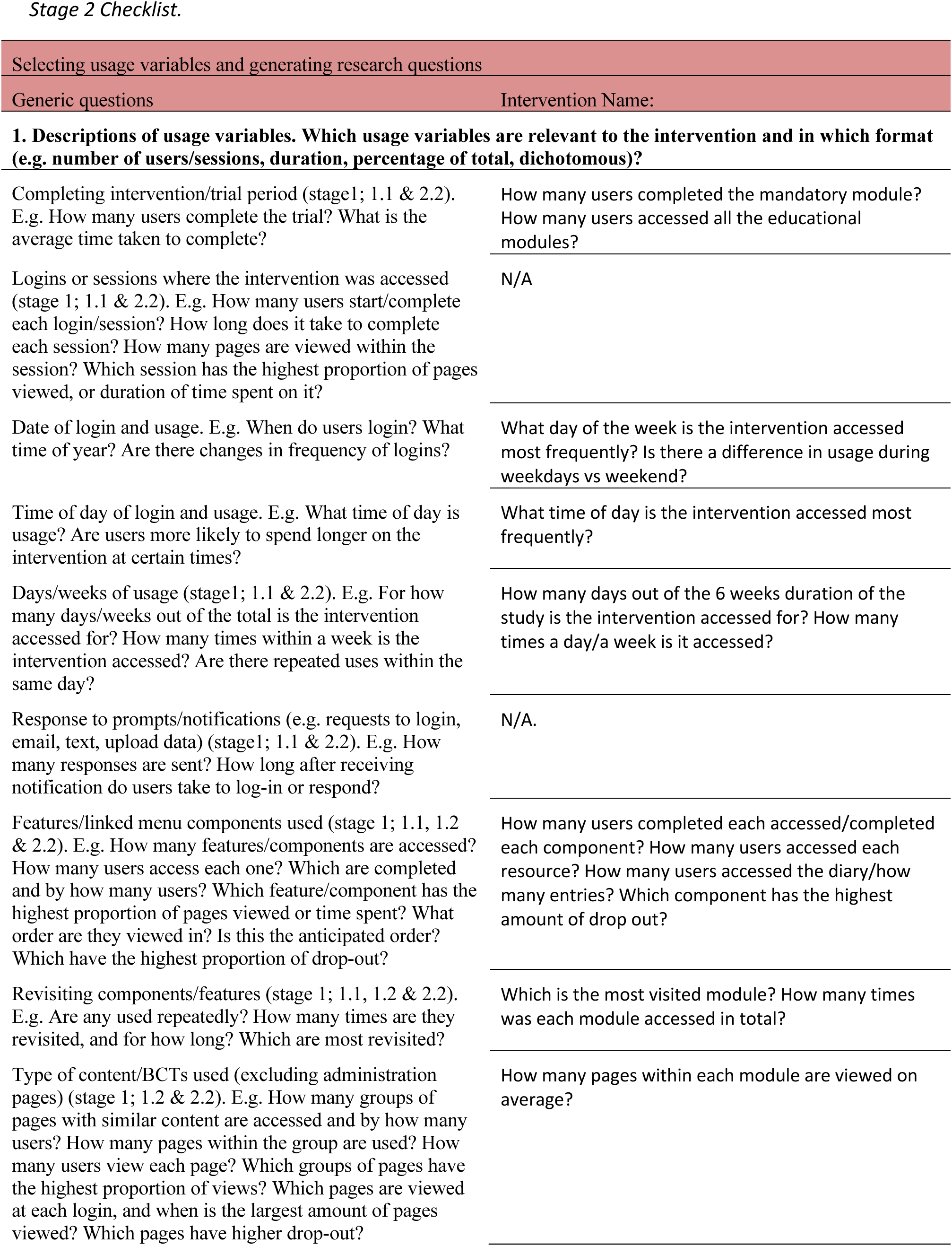
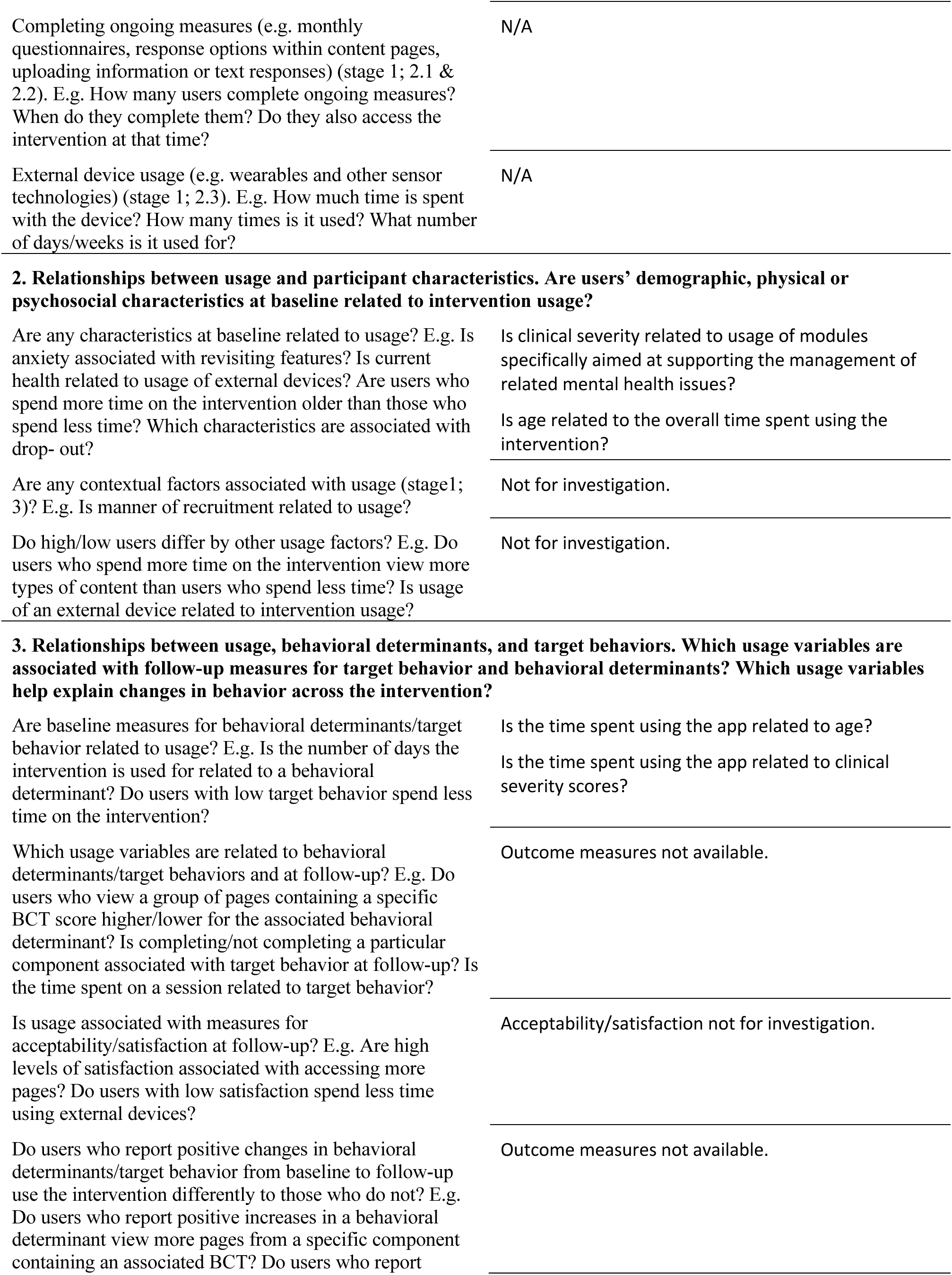
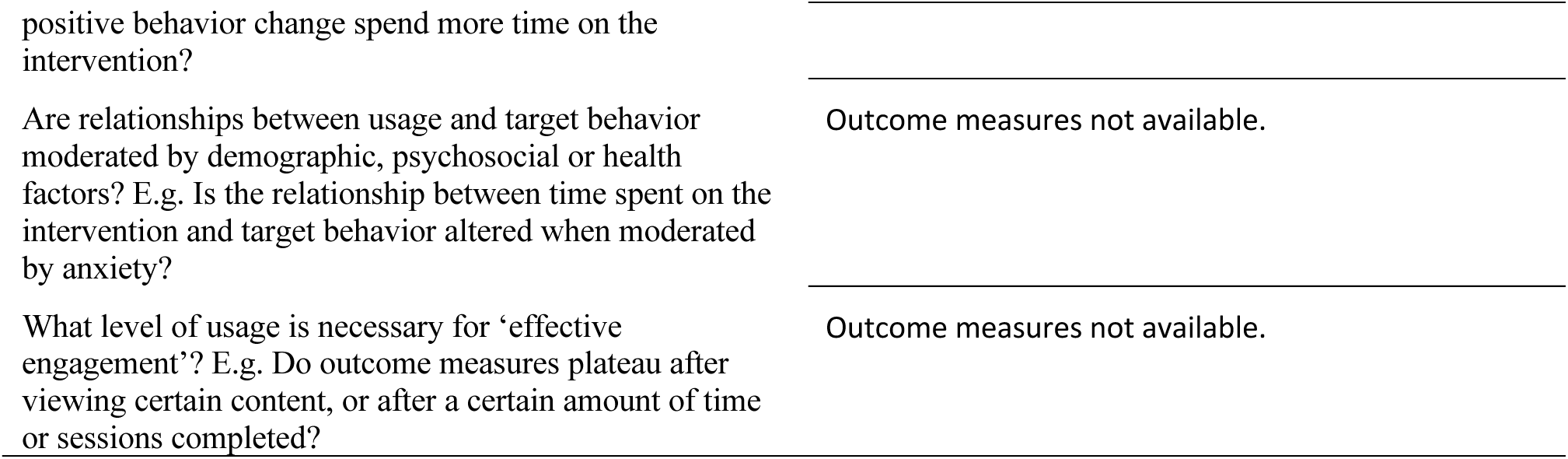
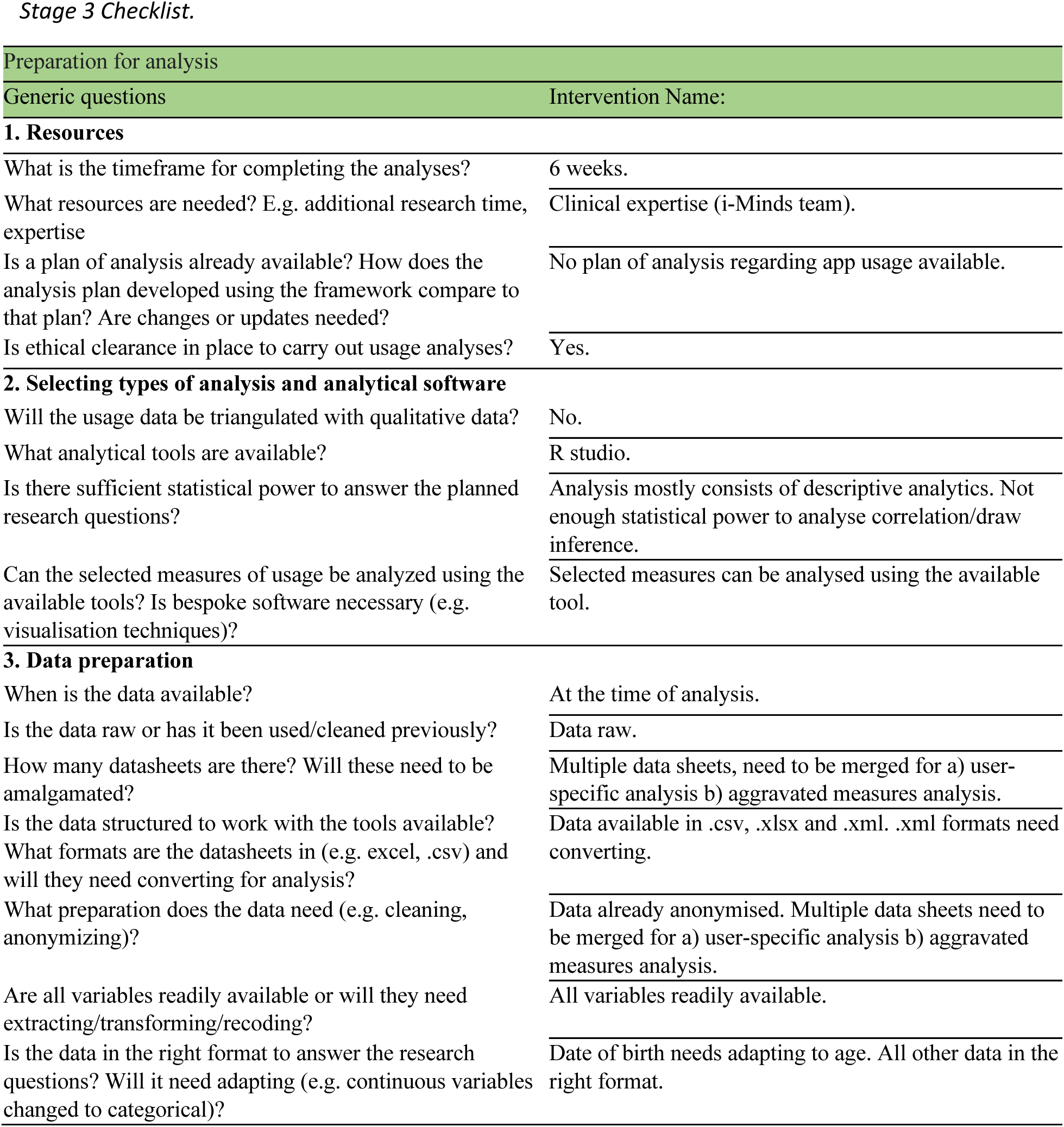
AMUsED checklists.

**Supplementary Figure 1.**
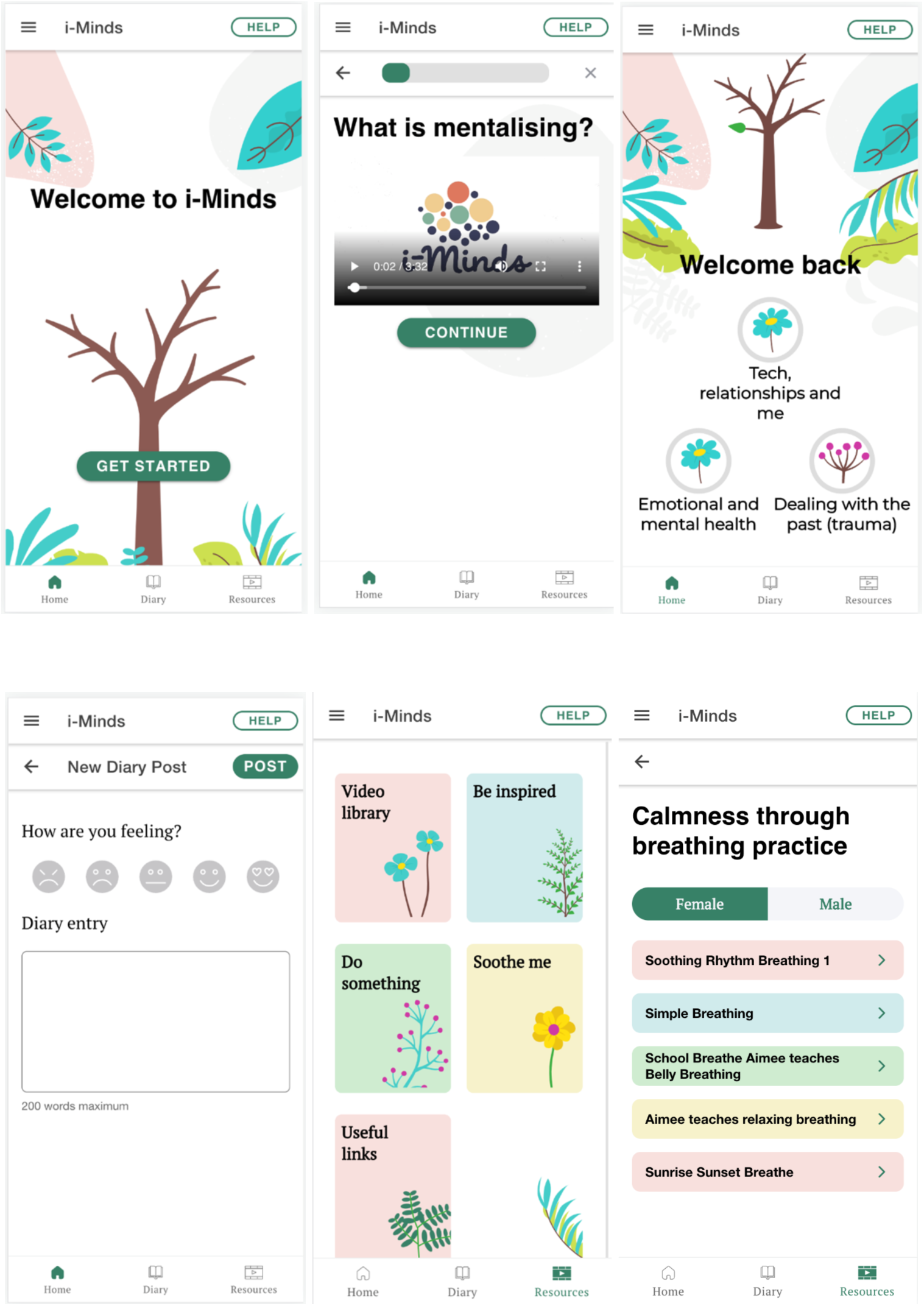
Screenshots of the i-Minds app.

**Supplementary Figure 2.**
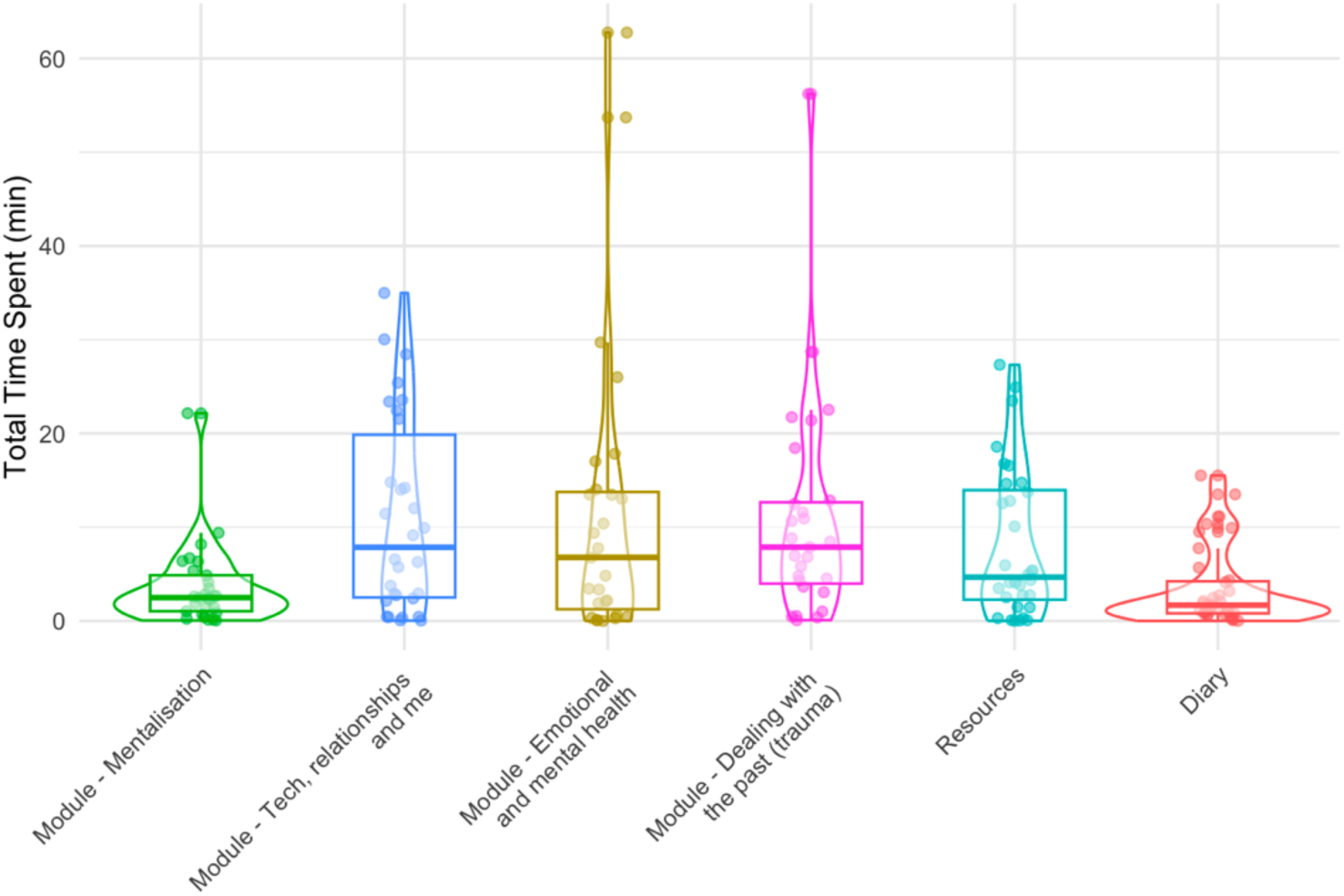
Total time spent using each app feature Scatter plot shows the total time spent on specific app contents of each participant (data points were jittered to reduce overlapping); box plot and violin plot show the distribution of the data points.

**Supplementary Figure 3.**
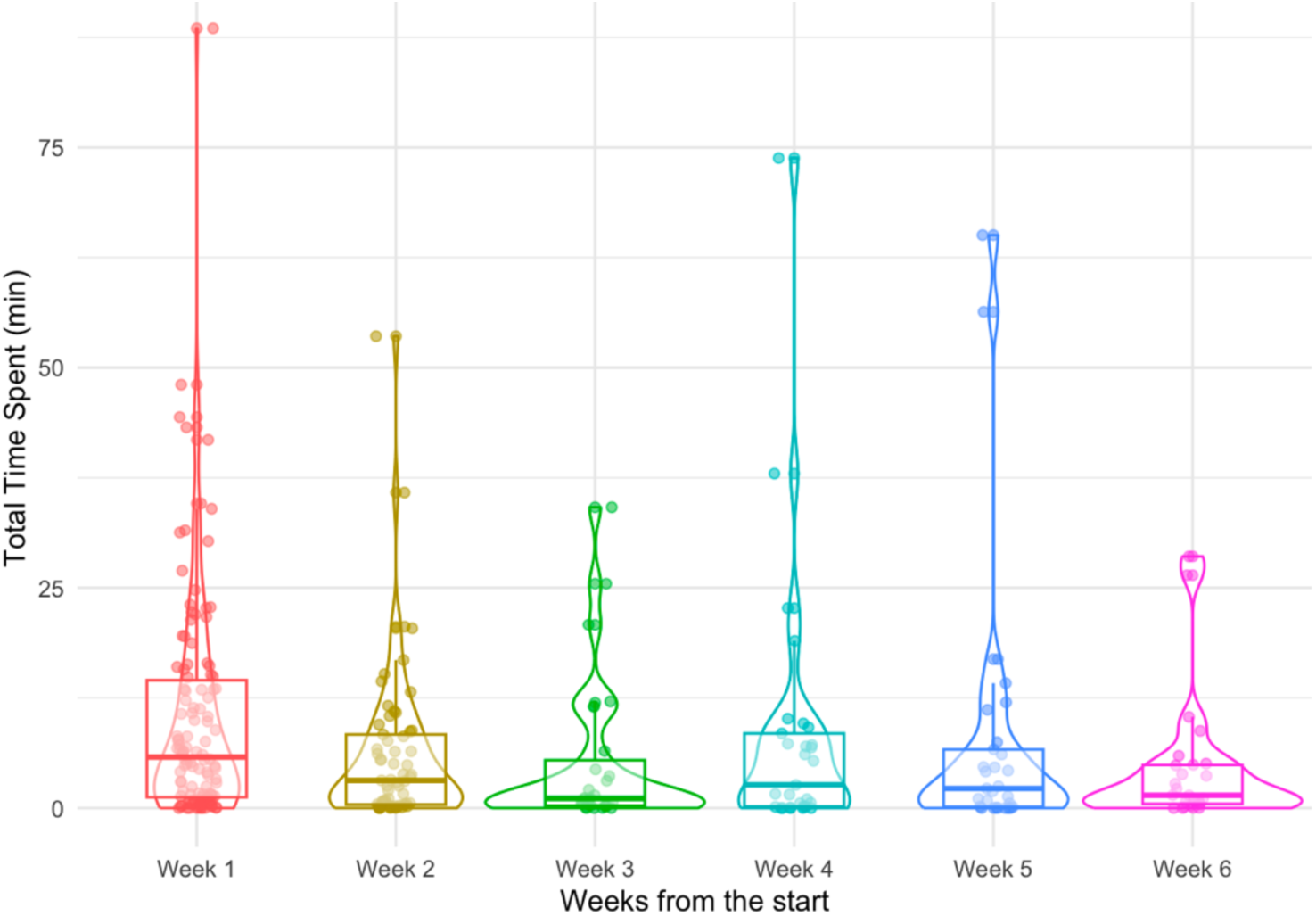
Daily total time spent using the app over 6weeks Scatter plot shows the daily total time spent of each participant in each week (data points were jittered to reduce overlapping); box plot and violin plot show the distribution of the data points.

